# Fertility care among people with primary ciliary dyskinesia

**DOI:** 10.1101/2023.07.04.23292228

**Authors:** Leonie D Schreck, Myrofora Goutaki, Philippa Jörger, Katie Dexter, Michele Manion, Sophie Christin-Maitre, Bernard Maitre, COVID-PCD patient advisory group, Claudia E Kuehni, Eva SL Pedersen

**Affiliations:** Institute of Social and Preventive Medicine, University of Bern, Bern, Switzerland; Graduate School for Health Sciences, University of Bern, Bern, Switzerland; Division of Paediatric Respiratory Medicine and Allergology, Department of Paediatrics, Inselspital, University Hospital, University of Bern, Bern, Switzerland; PCD Support UK, London, United Kingdom; PCD Foundation, Minneapolis, USA; Sorbonne University, INSERM UMR 933, 75012 Paris, France; Univ Paris Est-Créteil, Faculté de Santé, INSERM, IMRB, Créteil, France; Pulmonary Department, Centre Hospitalier Intercommunal de Créteil, France

## Abstract

**Introduction:** Fertility care is important for people living with primary ciliary dyskinesia (PCD) who are at increased risk of fertility problems. We investigated fertility care in an international participatory study.

**Methods:** Participants of the COVID-PCD study completed an online questionnaire addressing fertility issues. We used logistic regression to study factors associated with fertility specialist visits.

**Results:** Among 384 respondents (response rate 53%), 266 were adults [median age 44 years, interquartile range (IQR) 33–54), 68% female], 16 adolescents, and 102 parents of children with PCD. Half adult participants (128; 48%) received care from fertility specialists at a median age of 30 years (IQR 27–33)—a median of 10 years after PCD diagnosis. Fertility specialist visits were reported more often by adults with pregnancy attempts [odds ratio (OR) 9.1, 95% confidence interval (CI) 3.8–23.6] and among people who reported fertility as important for them (OR 5.9, 95% CI 2.6–14.6) and less often by females (OR 0.4, 95% CI 0.2–0.8). Only 56% of participants who talked with healthcare professionals about fertility were satisfied with information they received. They expressed needs for more comprehensive fertility information and reported dissatisfaction with physician knowledge about PCD and fertility.

**Conclusion:** People with PCD are inconsistently referred to fertility specialists. We recommend care from fertility specialists become standard in routine PCD care, and that PCD physicians provide initial fertility information either at diagnosis or no later than transition to adult care.

## Introduction

Fertility care is important for people living with primary ciliary dyskinesia (PCD) who are at increased risk of fertility problems. PCD is a rare genetic disease which affects the function and structure of motile cilia. Motile cilia are found in the respiratory tract, fallopian tubes, and efferent ductules ^1, 2^. In addition, sperm flagella share common structures with motile cilia ^3^. In females, it was hypothesized the abnormal motion of cilia in fallopian tubes possibly leads to infertility and more ectopic pregnancies ^1, 4, 5^. Male infertility is likely caused by reduced sperm count in ejaculate from altered motility of cilia in the efferent ductules or by dysmotile sperm ^2, 3, 6^. However, the extent of fertility problems—especially among females—is unknown ^7^.

Fertility care is defined as interventions that include fertility awareness, support and fertility management ^8^. Patients with PCD would benefit from fertility care as it provides information for making informed decisions about reproductive health. If needed, assisted reproduction interventions, such as *in vitro* fertilization with or without intracytoplasmic sperm injections, could be offered early on to increase chances of pregnancy. However, no clear guidelines currently exist for integrating fertility care into routine PCD care. Recommendations about fertility care are scarce and imprecise ^9–15^. It is unclear how fertility care is implemented into routine PCD care, what information is provided and when, and whether people with PCD are satisfied with fertility information they receive. For cystic fibrosis (CF)—another rare genetic lung disease—fertility issues are also present among most males and some females. Studies about CF and reproductive health suggest adults, adolescents, and parents of children with CF know about the possibility of fertility problems, yet lack important CF-specific reproductive health information ^16–20^. CF and reproductive health studies share clear recommendations for standardized education about reproductive health ^16, 21^.

Using data from the largest study worldwide collecting information directly from people with PCD (COVID-PCD), we studied fertility care among people with PCD, including their sources of and factors for referrals to fertility specialists and satisfaction with fertility information received.

## Methods

### Study design and ethics

We obtained study data from a cross-sectional questionnaire about fertility nested within the COVID-PCD study. COVID-PCD is an international participatory cohort study that collects information directly from people with PCD (clinicaltrials.gov: NCT04602481). A detailed study protocol has been published ^22^. In brief, the COVID-PCD study was set up in 2020 at the University of Bern, Switzerland in collaboration with international PCD patient support groups to follow people with PCD through the pandemic and answer other PCD-related research questions. The COVID-PCD study enrolls participants of any age with PCD from all over the world with suspected or confirmed PCD diagnoses. The study is online, anonymous, and longitudinal. Questionnaires are available in five languages (English, French, German, Italian, and Spanish).

The cantonal ethics committee of Bern approved the study (study ID: 2020-00830). We obtained informed consent either from participants or parents of participants younger than age 14. We report according to strengthening the reporting of observational studies in epidemiology (STROBE) recommendations ^23^.

### Study procedures

Participants registered via the study website (www.covid19pcd.ispm.ch) and received a baseline questionnaire, then weekly follow-up questionnaires. We sent a fertility questionnaire by email on July 12, 2022 to all participants enrolled in the COVID-PCD study. We sent up to three reminders if participants did not yet complete the questionnaire. We collected responses until March 8, 2023. All data was entered directly in a web-based database using the Research Electronic Data Capture (REDCap) platform ^24^ hosted by the Swiss medical registries and data linkage center (SwissRDL) at the University of Bern.

### Fertility questionnaire

We developed the fertility questionnaire based on existing literature, and we discussed it with fertility specialists and patient representatives from PCD support groups. We include questions and answer categories in Supplementary Table S1. We developed separate questionnaires for adults older than 18 years, adolescents ages 14–17 years, and parents of children with PCD younger than 14 years. Native speaking members from patient support groups or our research team translated questionnaires; two research team members independently checked translations.

### Information on fertility care characteristics

In the adult questionnaire, we asked participants if ever referred to fertility specialists; if so, at what age and who provided referrals. We calculated time until fertility specialist visit as the difference between age at PCD diagnosis (or age 18 for people diagnosed during childhood) and age at fertility specialist visit. We excluded adults who were diagnosed with PCD only after fertility specialist visits or with unreported ages at PCD diagnosis or at fertility specialist visit.

We asked all participants (adults, adolescents, and parents) their opinions about the need for fertility care, their fertility information seeking behaviors, and sources of fertility information they used. Among participants who previously talked with healthcare professional about fertility, we asked about what topics were discussed and whether they were satisfied with fertility information they received. We asked participants who reported they were either partly satisfied or unsatisfied with provided fertility information to pinpoint information gaps.

We studied how fertility care from fertility specialists differed by age and country. For adults, age at survey, age at diagnosis, and age at fertility specialist visit were categorized as either 18–30 years, 31–45 years, or older than 45 years. We chose age groups according to physiological female fertility changes with peak reproductive years between late teens and late 20s; declining fertility starting by age 30 and by age 45, fertility declining so much pregnancy without medical assistance is unlikely for most females ^25^. For simplicity, we used the same age groups for males. We categorized country of residence. We combined Canada and the United States as North America because their PCD care is organized in a network. We included England, Northern Ireland, Scotland, and Wales as the United Kingdom. We grouped countries with fewer than 25 participants into either “other European countries” or “other non-European countries” (Supplementary Table S2).

## Data analysis

We described fertility care separately for females and males. We compared time from diagnosis (or age 18 for people diagnosed during childhood) until fertility specialist visit between females and males using Mann Whitney U test. We studied factors associated with fertility specialist visits using multivariable logistic regression with sex, age at survey, age at PCD diagnosis, pregnancy attempt, and self-assessed importance of fertility as explanatory variables. For the regression analysis, we excluded people with missing values for age at PCD diagnosis and without information on pregnancy attempts. Each variable had <5% missing data. We used R version 4.2.0 for all analyses ^26^.

## Results

### Study population

In total, 384 of 723 COVID-PCD participants (response rate 53%) completed the fertility questionnaire (Supplementary Table S3). Of these, 266 were adults (69%) with a median age of 44 years [interquartile range (IQR) 33–54]; 16 adolescents (4%), and 102 parents of children with PCD (27%). Two-thirds of adult participants were female (181; 68%). Most participants were from North America (71; 18%), the United Kingdom (66; 17%), and Germany (62; 16%). Compared with non-participants, people who completed the fertility questionnaire were older and more often living in Germany, Switzerland, Australia, and other European countries (Supplementary Table S3).

### Fertility care among adults

Half of adult participants (128; 48%) reported receiving fertility care from fertility specialists before completing the survey (Table 1) with a higher proportion of males (47; 55%) than females (81; 45%). The median age at fertility specialist visit was 30 years (IQR 27–33). Only 15 (12%) participants were referred by their PCD physicians, while most participants either reported referrals by non-PCD physicians (46; 36%) or organized appointments themselves (43; 34%) (Table 1). The median time between PCD diagnosis (or age 18 for people diagnosed during childhood) and fertility specialist visit was 10 years, and higher among females (11 years, IQR 7–13) than males (7.5 years, IQR 0–14) (p-value 0.06).

**Table 1:** Frequency of fertility specialist visits, age at visit, mode of referral, and time until fertility specialist visit among adult females (n = 181) and males (n = 85) with PCD (COVID-PCD study).

| Adult participants who completed the fertility questionnaire | Overall<br>n = 266<br>n (%) | Female<br>n = 181<br>n (%) | Male<br>n = 85<br>n (%) |
| --- | --- | --- | --- |
| **Have you ever visited a fertility specialist?** |  |  |  |
| Yes | 128 (48) | 81 (45) | 47 (55) |
| No, but I have an appointment | 6 (2) | 5 (3) | 1 (1) |
| No, but I would like to | 14 (5) | 12 (7) | 2 (2) |
| No | 118 (44) | 83 (46) | 35 (41) |
| **Adult participants who visited fertility specialists** |  |  |  |
| **Age at fertility specialist visit** |  |  |  |
| Median (IQR) | 30 (27–33) | 29 (27–32) | 30 (28–35) |
| 18–30 y | 74 (58) | 50 (62) | 24 (51) |
| 31–45 y | 53 (41) | 30 (37) | 23 (49) |
| Missing | 1 (1) | 1 (1) | 0 (0) |
| Referral by PCD physician | 15 (12) | 5 (6) | 10 (21) |
| Referral by non-PCD physician | 46 (36) | 29 (36) | 17 (36) |
| Participant asked for referral | 19 (15) | 14 (17) | 5 (11) |
| Participant organized appointment | 43 (34) | 28 (35) | 15 (32) |
| Other<sup>a</sup> | 2 (2) | 2 (2) | 0 (0) |
| Missing | 3 (2) | 3 (4) | 0 (0) |
| **Time until fertility specialist visit<sup>b</sup>** |  |  |  |
| Years, median (IQR) | 10 (6–13) | 11 (7–13) | 7.5 (0–14) |

<sup>a</sup>Other answers included friend (n = 1), not specified (n = 1). <sup>b</sup>Calculated as the difference between the age at PCD diagnosis (or age 18 for people diagnosed during childhood) and the age at fertility specialist visit. We excluded participants when PCD was diagnosed only after fertility specialist visit (female n = 10; male n = 15) and when age at diagnosis (female n = 9) or age at fertility specialist visit (female n = 1) were unreported. All characteristics are presented as n and column %, unless otherwise stated. Abbreviations: IQR, interquartile range. PCD, primary ciliary dyskinesia. y, years.

Adults who received fertility care from fertility specialists were less often female [odds ratio (OR) 0.4, 95% confidence interval (CI) 0.2–0.8], more often reported prior pregnancy attempts (OR 9.1, 95% CI 3.8–23.6) and fertility as an important issue (OR 5.9, 95% CI 2.6–14.6) (Table 2). The proportion of people who reported fertility care from fertility specialists differed between countries with the highest in France (11; 69%) and lowest in Italy (7; 35%).

**Table 2:**
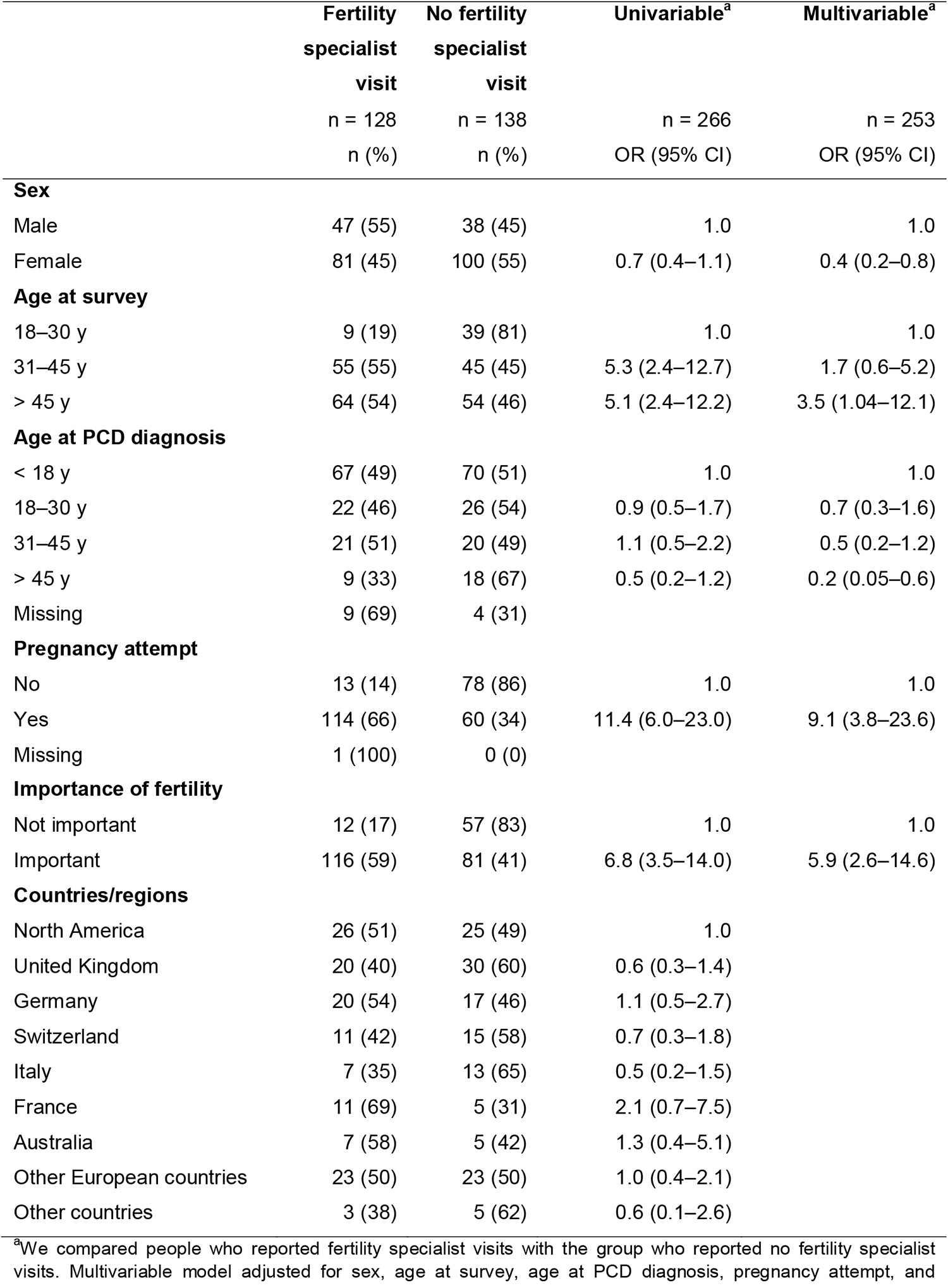
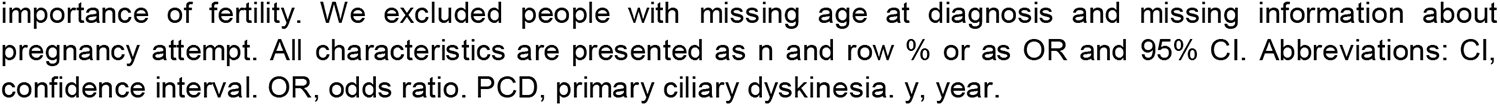
Characteristics of adult participants and predictors of fertility specialist visit (COVID-PCD study, n = 266).

|  | Fertility specialist visit<br>n = 128<br>n (%) | No fertility specialist visit<br>n = 138<br>n (%) | Univariable<sup>a</sup><br>n = 266<br>OR (95% CI) | Multivariable<sup>a</sup><br>n = 253<br>OR (95% CI) |
| --- | --- | --- | --- | --- |
| **Sex** |  |  |  |  |
| Male | 47 (55) | 38 (45) | 1.0 | 1.0 |
| Female | 81 (45) | 100 (55) | 0.7 (0.4–1.1) | 0.4 (0.2–0.8) |
| **Age at survey** |  |  |  |  |
| 18–30 y | 9 (19) | 39 (81) | 1.0 | 1.0 |
| 31–45 y | 55 (55) | 45 (45) | 5.3 (2.4–12.7) | 1.7 (0.6–5.2) |
| > 45 y | 64 (54) | 54 (46) | 5.1 (2.4–12.2) | 3.5 (1.04–12.1) |
| **Age at PCD diagnosis** |  |  |  |  |
| < 18 y | 67 (49) | 70 (51) | 1.0 | 1.0 |
| 18–30 y | 22 (46) | 26 (54) | 0.9 (0.5–1.7) | 0.7 (0.3–1.6) |
| 31–45 y | 21 (51) | 20 (49) | 1.1 (0.5–2.2) | 0.5 (0.2–1.2) |
| > 45 y | 9 (33) | 18 (67) | 0.5 (0.2–1.2) | 0.2 (0.05–0.6) |
| Missing | 9 (69) | 4 (31) |  |  |
| **Pregnancy attempt** |  |  |  |  |
| No | 13 (14) | 78 (86) | 1.0 | 1.0 |
| Yes | 114 (66) | 60 (34) | 11.4 (6.0–23.0) | 9.1 (3.8–23.6) |
| Missing | 1 (100) | 0 (0) |  |  |
| **Importance of fertility** |  |  |  |  |
| Not important | 12 (17) | 57 (83) | 1.0 | 1.0 |
| Important | 116 (59) | 81 (41) | 6.8 (3.5–14.0) | 5.9 (2.6–14.6) |
| **Countries/regions** |  |  |  |  |
| North America | 26 (51) | 25 (49) | 1.0 |  |
| United Kingdom | 20 (40) | 30 (60) | 0.6 (0.3–1.4) |  |
| Germany | 20 (54) | 17 (46) | 1.1 (0.5–2.7) |  |
| Switzerland | 11 (42) | 15 (58) | 0.7 (0.3–1.8) |  |
| Italy | 7 (35) | 13 (65) | 0.5 (0.2–1.5) |  |
| France | 11 (69) | 5 (31) | 2.1 (0.7–7.5) |  |
| Australia | 7 (58) | 5 (42) | 1.3 (0.4–5.1) |  |
| Other European countries | 23 (50) | 23 (50) | 1.0 (0.4–2.1) |  |
| Other countries | 3 (38) | 5 (62) | 0.6 (0.1–2.6) |  |

<sup>a</sup>We compared people who reported fertility specialist visits with the group who reported no fertility specialist visits. Multivariable model adjusted for sex, age at survey, age at PCD diagnosis, pregnancy attempt, and
importance of fertility. We excluded people with missing age at diagnosis and missing information about pregnancy attempt. All characteristics are presented as n and row % or as OR and 95% CI. Abbreviations: CI, confidence interval. OR, odds ratio. PCD, primary ciliary dyskinesia. y, year.

Most participants (370; 96%) felt people with PCD should be referred for fertility care by fertility specialists (Table 3), yet opinions about the timing of fertility care differed. More than one-third (144; 38%) reported everyone should be referred; many (102; 27%) thought only people who want children should be referred; some (78; 20%) felt only adults and few (37; 10%) reported only people with unsuccessful pregnancy attempts should be referred.

**Table 3:** Participant opinions on fertility care and their sources of fertility information (COVID-PCD study, n = 384).

| Participants who completed the fertility questionnaire | Overall | Female | Male | Adolescent | Parent of children |
| --- | --- | --- | --- | --- | --- |
|  | n = 384<br>n (%) | n = 181<br>n (%) | n = 85<br>n (%) | n = 16<br>n (%) | n = 102<br>n (%) |
| **Who should be referred to fertility specialists?** |  |  |  |  |  |
| Everyone | 144 (38) | 66 (36) | 37 (44) | 7 (44) | 34 (33) |
| Only adults | 78 (20) | 25 (14) | 14 (16) | 3 (19) | 36 (35) |
| Only people who want children | 102 (27) | 54 (30) | 25 (29) | 5 (31) | 18 (18) |
| Only people with unsuccessful pregnancy attempts | 37 (10) | 24 (13) | 5 (6) | 0 (0) | 8 (8) |
| Other<sup>a</sup> | 9 (2) | 2 (1) | 1 (1) | 1 (6) | 5 (5) |
| Nobody | 14 (4) | 10 (6) | 3 (4) | 0 (0) | 1 (1) |
| **I searched for information about fertility** | 277 (72) | 130 (72) | 65 (76) | 8 (50) | 74 (73) |
| Missing | 1 (0) | 1 (1) | 0 (0) | 0 (0) | 0 (0) |
| **Where did you search for information?** | **n = 277** | **n = 130** | **n = 65** | **n = 8** | **n = 74** |
| PCD network websites | 119 (43) | 53 (41) | 20 (31) | 4 (50) | 42 (57) |
| Scientific articles | 92 (33) | 47 (36) | 20 (31) | 1 (13) | 24 (32) |
| Patient support groups/<br>patients with PCD | 88 (32) | 43 (33) | 13 (20) | 1 (13) | 31 (42) |
| Other websites | 87 (31) | 41 (32) | 13 (20) | 4 (50) | 29 (39) |
| Social media | 41 (15) | 21 (16) | 7 (11) | 1 (13) | 12 (16) |
| Friends/family | 17 (6) | 11 (8) | 4 (6) | 2 (25) | 0 (0) |
| Books | 6 (2) | 3 (2) | 2 (3) | 0 (0) | 1 (1) |
| **I talked about fertility with a healthcare professional** | 203 (53) | 108 (60) | 61 (72) | 2 (13) | 32 (31) |

<sup>a</sup>Other answers included mid-late teens/reproductive age (n = 3), only if they want to (n = 4), children at an age determined by parent/guardian (n = 1), not specified (n = 1). Abbreviation: PCD, primary ciliary dyskinesia. All characteristics are presented as n and column %.

### Fertility information among all participants

Most participants (277; 72%) searched for information about fertility (Table 3). They mostly searched multidisciplinary PCD network websites (119; 43%) or scientific articles (92; 33%). Around half (203; 53%) of all participants talked about fertility with healthcare professionals involved in their PCD care prior to the study, with adult females (108; 60%) and adult males (61; 72%) more often than parents of children (32; 31%) and adolescents (2; 13%) with PCD (Table 3). Participants reported most healthcare professionals mentioned people with PCD may have problems with fertility (females: 97; 90%, males: 53; 87%, parents: 30; 94%) (Figure 1). More than half of adult participants received information about the possibility of fertility treatments (females: 67; 62%, males: 33; 54%) and one-quarter (females: 31; 29%, males: 16; 26%) were informed about fertility awareness factors other than PCD influencing fertility, such as advanced age or lifestyle factors. Overall, half of participants (111; 56%) were satisfied with information from healthcare professionals, ranging from fewer than half of females (49; 45%) to three-quarters of males (43; 72%) (Figure 2). Participants who reported dissatisfaction with fertility information noted information gaps, such as detailed information about effects from PCD on fertility (18; 20%), effects on female fertility specifically (8; 9%) or information and advice on fertility treatments (9; 10%). They reported dissatisfaction with physician knowledge about PCD and fertility (16; 18%). Eight participants reported fertility was not yet relevant because the person diagnosed with PCD was young; and five reported long ago PCD-fertility discussions with little known about PCD at the time of the conversation (Table 4).

**Figure 1:**
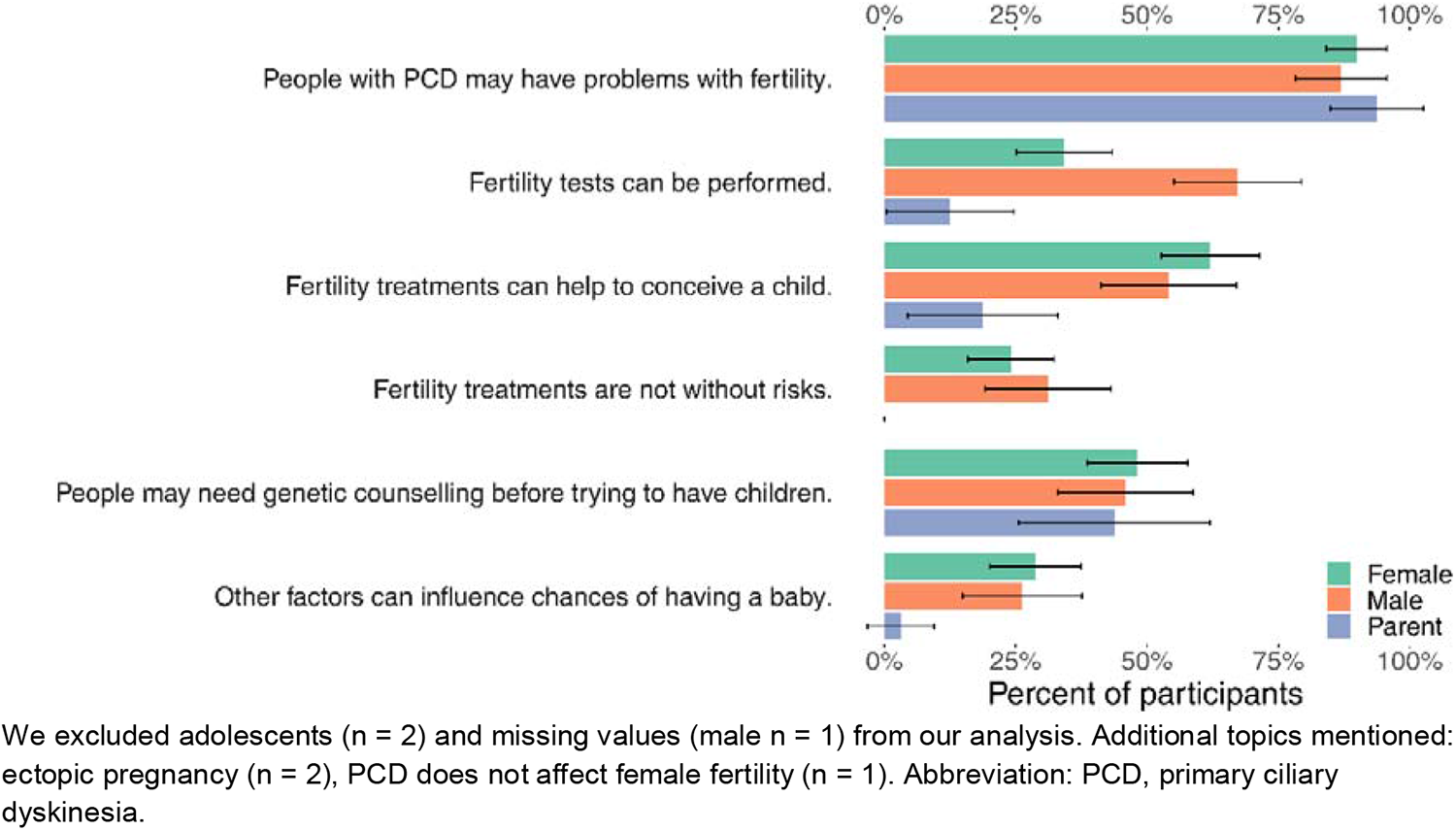
Topics covered in fertility discussions with the healthcare professional among females (n = 108), males (n = 60) and parents of children with PCD (n = 32).

**Figure 2:**
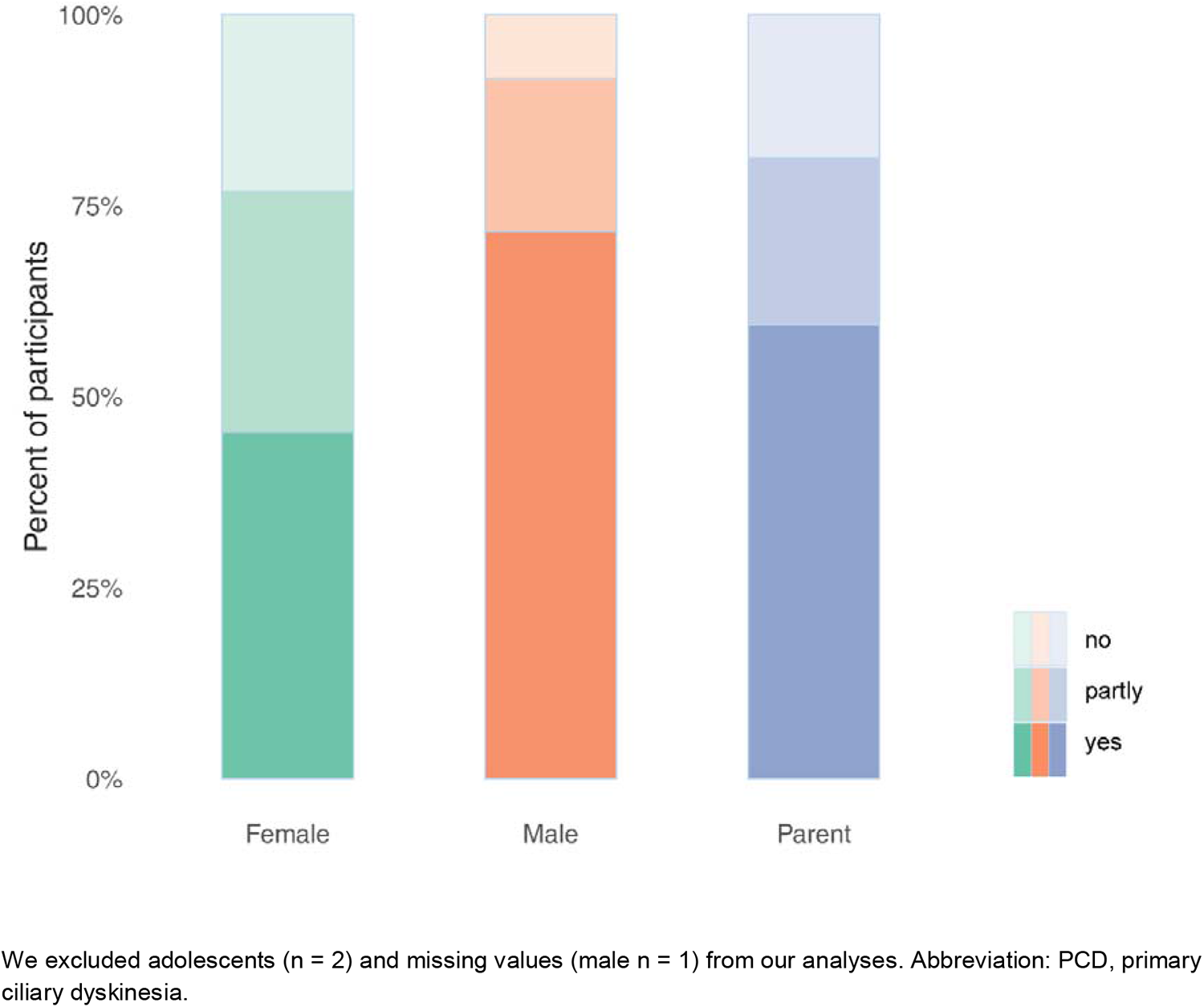
Satisfaction with information provided by fertility specialists among females (n = 108), males (n = 60), and parents of children with PCD (n = 32) (COVID-PCD study).

**Table 4:** Fertility information gaps identified and other reasons for dissatisfaction among people who discussed fertility care with healthcare professionals (n = 89) (COVID-PCD study).

| Participants who were dissatisfied with information about fertility from healthcare professionals (n = 89) | n (%) |
| --- | --- |
| **Fertility information gaps** |  |
| Detailed information about effects from PCD on fertility | 18 (20) |
| Specific information |  |
| – Effects of PCD on female fertility | 8 (9) |
| – Chances of pregnancy without medical assistance | 5 (6) |
| – Differences between fertility problems and sterility | 2 (2) |
| – Risks of ectopic pregnancy | 2 (2) |
| – Information about pregnancy and PCD | 2 (2) |
| – Risk of inherited PCD | 1 (1) |
| – Associations of fertility with genetic defects | 1 (1) |
| Follow-up care |  |
| – Information and advice on fertility treatments | 9 (10) |
| – Information on fertility tests | 2 (2) |
| – Offering genetic counselling | 1 (1) |
| – Follow-up care when ready for pregnancy | 1 (1) |
| – Sources of further information | 1 (1) |
| – Support for adoption | 1 (1) |
| **Other reasons for dissatisfaction** |  |
| Physician knowledge of PCD and fertility | 16 (18) |
| Fertility was not yet relevant because person diagnosed with PCD was young | 8 (9) |
| Little known about PCD at the time of the conversation | 5 (6) |
| Emotional component, physician empathy | 4 (4) |
| PCD diagnosed after the conversation | 2 (2) |
| Lack of scientific evidence from studies | 2 (2) |
| Information about fertility not provided earlier | 1 (1) |

All characteristics are presented as n and column %. Abbreviation: PCD, primary ciliary dyskinesia.

### Discussion Summary of findings

In the first study on fertility care among people with PCD, we showed only 48% of adults with PCD reported ever visiting fertility specialists with a median delay of 10 years since diagnosis (or age 18 for people diagnosed during childhood). Only 12% were referred to fertility specialists by their PCD physician. Among participants with PCD who spoke with healthcare professional about fertility, only 56% reported satisfaction with provided information. They expressed needs for more comprehensive fertility information, such as detailed information about effects from PCD on fertility.

### Strengths and weaknesses

Using data from the largest study with patient-reported data in PCD, we collected information about fertility care and opinions from people with PCD. Above all, our results represent relevance for people with PCD. Since we conducted our study online and anonymously, we involved people normally with limited involvement in research from many countries around the world.

Our study also has limitations. People who talked with healthcare professionals about fertility long ago possibly found it difficult to recall conversation details, which could lead to recall bias. However, we did not ask detailed questions about fertility discussions; instead, we focused on general aspects and opinions. Therefore, we expected respondents to remember the main points discussed. Fertility is a sensitive topic and people with negative fertility care experiences possibly participate more as an opportunity to make themselves heard, which could lead to selection bias.

### Comparison with other studies

To our knowledge, no other study investigated fertility care among people with PCD. Review papers on managing PCD suggest including fertility topics in routine PCD care ^10, 15^. The PCD Foundation recommends including reproductive medicine “as clinically needed” ^9^; Werner et al. note “infertility is managed with adequate reproductive techniques” ^14^; and Lucas et al. suggest integrating fertility care at transition into adult care ^13^. For male infertility, two reviews promote including fertility care during the transition from pediatric to adult care ^27 3^. However, none explicitly counsel how and when fertility care should be included; how it should be organized; and what topics should be addressed ^7^. In CF research, recent reproductive health reviews address some of these questions and propose management strategies for healthcare professionals ^21, 28–31^. However, data on fertility care implementation show reproductive health needs are not proactively addressed among people with CF ^32^. Adults and adolescents with CF report inadequate communication on the topic from CF care providers ^17, 18^ and delayed introduction of reproductive health education ^16^. Only 30% of parents of children with CF were satisfied with their current knowledge of reproductive health ^20^. Results from our study confirm people with PCD face similar challenges in terms of fertility care and highlight important gaps in PCD care and existing recommendations.

### Interpretation of results

Young adults with PCD are not referred to fertility specialists as part of routine PCD care. Most seem only to be referred when experiencing fertility problems, and usually at their own request. Females are referred later than males, although an early visit would be even more important for them since their fertility declines earlier. This suggests fertility care is not satisfactorily integrated in routine PCD care and potential fertility problems are not proactively addressed before becoming relevant to people with PCD. Only one-quarter of participants were informed by healthcare professionals about other fertility awareness factors, such as age, influencing fertility; only half of participants were informed about the possibility of fertility treatments. Our results also suggest great dissatisfaction among people with PCD regarding their fertility care. Most participants expressed a need for more comprehensive information about fertility for people with PCD. This also reflects the general lack of knowledge regarding fertility issues among people with PCD—especially in females—and indicates a need for further research. Until this knowledge gap is filled, fertility care will have limitations.

PCD physicians may not recognize the need to address fertility as it is out of their area of expertise. Nevertheless, it is likely beneficial for PCD physicians to openly discuss potential fertility issues. Already at diagnosis, PCD physicians can provide information about potential impact from PCD on fertility and—no later than the transition to adult care—offer information about fertility awareness, such as age or timing of sexual intercourse. By integrating fertility care, physicians could mention ways to increase chances of pregnancy through fertility treatments and interventions, offer further fertility information, and organize referrals to fertility specialists if needed.

## Conclusion

People with PCD are inconsistently referred to fertility specialists and report difficulties obtaining satisfactory fertility information. We recommend care from fertility specialists become standard in routine PCD care, and PCD physicians provide initial fertility information either at diagnosis or no later than transition to adult care.

## Data Availability

All data produced in the present study are available upon reasonable request to the authors

## Acknowledgments

We thank all participants and their families, and we thank the PCD support groups and physicians who advertised the study. We thank our collaborators who helped set up the COVID-PCD study: Cristina Ardura, Yin Ting Lam, Christina Mallet, Helena Koppe, Dominique Rubi from the University of Bern and Jane S Lucas and Amanda Harris from the University Hospital Southampton. We thank Nathalie Massin and Lara Gonçalves Pissini for their help in the design of the fertility questionnaire. We thank Kristin Marie Bivens for her editorial work and guidance on our manuscript.

## Declarations

### Ethics approval and consent to participate

Bern Cantonal Ethics Committee (Kantonale Ethikkomission Bern) approved our study (Study ID: 2020-00830). Participants provide their informed consent to participate in the study at registration. Participation is anonymous. Participants can withdraw consent to participate at any time by contacting the study team.

### Availability of data and materials

COVID-PCD data is available upon reasonable request by contacting Claudia Kuehni.

### Authors’ contributions

LD Schreck, M Goutaki, CE Kuehni and ESL Pedersen made substantial contributions to the study concept and design. LD Schreck, M Goutaki, K Dexter, M Manion, S Christin-Maitre, B Maitre, Claudia E Kuehni and ESL Pedersen were involved in the design of the fertility questionnaire. LD Schreck analyzed data and drafted the manuscript. LD Schreck, ESL Pedersen, P Jörger, K Dexter, M Manion, S Christin-Maitre, B Maitre, M Goutaki, and CE Kuehni critically revised and approved the manuscript.

## Supplementary Material

**Supplementary Table S1:** Formulation of questions from the English adult baseline questionnaire and the English female and male fertility questionnaires from the COVID-PCD study.

| Question | Answer category |
| --- | --- |
| **Baseline Questionnaire** |  |
| What is your sex? | Male; Female; Other |
| Which year were you born? | *text (integer)* |
| Which country do you live in? | List of all countries worldwide |
| Which other country? | *text* |
| Which year were you diagnosed with PCD? | *text (integer)* |
| How old were you, when you were diagnosed with PCD? | *text (integer)* |
| **Fertility questionnaire** |  |
| Have you ever searched for information about fertility in people with PCD? | No; Yes |
| Where did you get your information from? (Tick all that apply) | Patient support groups;<br>Social media (e.g. Facebook, Twitter);<br>Websites from national or international PCD networks (e.g. BEAT-PCD);<br>Other websites;<br>Books;<br>Scientific articles;<br>Friends/family;<br>I asked my doctor;<br>Other |
| Please specify where you got your information from | *text* |
| Is fertility an important issue for you? | No; Yes |
| Have you ever talked to a health care professional about fertility and fertility-related problems of people with PCD? | No; Yes |
| What topics were covered in your discussion with the health care professional? (Tick all that apply) | People with PCD may have problems with fertility.;<br>PCD is a genetic disease and people may need counselling before trying to have children (genetics counselling).;<br>Fertility tests can be performed.;;<br>Fertility treatments can help to conceive a |
|  | child.;<br>Fertility treatments are not without risks.;<br>Other factors can influence chances of having a baby (e. g. timing of sexual intercourse, age, tobacco smoking).;<br>Other |
| Please specify what other topics were covered | *text* |
| Were you satisfied with the information about fertility provided by the health care professional? | No; Partly; Yes |
| What information was missing? | *text* |
| In your opinion, should every person with PCD receive fertility counselling from a fertility expert (= a doctor who is specialized in fertility)? | No;<br>Yes, everybody;<br>Yes, but only adults;<br>Yes, but only if they want children;<br>Yes, but only if they tried to get pregnant unsuccessfully;<br>Yes, other |
| Please specify who should receive fertility counselling | *text* |
| Have you ever seen a fertility expert (= a doctor who is specialized in fertility)? | No;<br>No, but I have an appointment / I am waiting to be referred;<br>No, but I would like to;<br>Yes |
| How old were you when you first saw a fertility expert? If you cannot remember the exact age, please give an approximate age. | *text (integer)* |
| Who referred you to the fertility expert? | I was referred by my PCD physician.;<br>I was referred by a doctor other than my PCD physician (e.g. my general practitioner).;<br>I had to ask for a referral.;<br>I had to organize it myself.;<br>Other |
| Please specify who referred you to the fertility expert | *text* |
| Have you ever tried to become pregnant?<br>Have you ever tried to father a child? | No;<br>Yes, but it did not work / we are still trying;<br>Yes, it worked |

**Supplementary Table S2:** Countries of residence of COVID-PCD participants with fewer than 25 enrolled participants who completed the fertility questionnaire, grouped as either “other European countries” or “other non-European countries”.

| Country | Number of participants |
| --- | --- |
| **Other European countries** | **67** |
| Austria | 4 |
| Belgium | 6 |
| Croatia | 1 |
| Cyprus | 2 |
| Czech Republic | 1 |
| Denmark | 5 |
| Georgia | 2 |
| Greece | 1 |
| Hungary | 1 |
| Ireland | 7 |
| Jersey | 1 |
| Netherlands | 12 |
| Norway | 8 |
| Poland | 3 |
| Portugal | 2 |
| Spain | 8 |
| Sweden | 3 |
| **Other non-European countries** | **9** |
| Brazil | 1 |
| Cameroon | 1 |
| Hong Kong | 1 |
| Israel | 2 |
| Panama | 1 |
| Puerto Rico | 1 |
| Saudi Arabia | 1 |
| South Africa | 1 |

**Supplementary Table S3:** Characteristics of participants who did or did not complete the fertility questionnaire (n = 723).

|  | Completed<br>questionnaire<br>n (%)<br>n = 384 | Not completed<br>questionnaire<br>n (%)<br>n = 339 |
| --- | --- | --- |
| **Questionnaire groups** |  |  |
| Adults | 266 (55) | 216 (45) |
| Adolescents | 16 (48) | 17 (52) |
| Parents | 102 (49) | 106 (51) |
| **Age at survey** |  |  |
| Adults, median (IQR; range) | 44 (33-54; 19-87) | 36 (26-47; 19-76) |
| 18-30 y | 48 (38) | 79 (62) |
| 31-45 y | 100 (56) | 78 (44) |
| > 45 y | 118 (67) | 59 (33) |
| Adolescents, median (IQR; range) | 17 (16-17; 15-18) | 17 (17-18; 16-18) |
| Children, median (IQR; range) | 9 (6-13; 2-15) | 9 (7-12; 3-15) |
| **Sex, adults** |  |  |
| Female | 181 (56) | 141 (44) |
| Male | 85 (54) | 73 (46) |
| Other | 0 (0) | 2 (100) |
| **Countries** |  |  |
| North America | 71 (46) | 85 (54) |
| United Kingdom | 66 (45) | 80 (55) |
| Germany | 62 (63) | 37 (37) |
| Switzerland | 36 (78) | 10 (22) |
| Italy | 29 (57) | 22 (43) |
| France | 23 (53) | 20 (47) |
| Australia | 21 (64) | 12 (36) |
| Other European countries | 67 (61) | 43 (39) |
| Other countries | 9 (23) | 30 (77) |

All characteristics are presented as n and row %, unless otherwise stated. Abbreviations: IQR, interquartile range. y, years.

